# The evolutionary genetics of polygenic diseases with the largest global burden in mortality rates

**DOI:** 10.1101/2021.12.10.21267630

**Authors:** Ujani Hazra, Joseph Lachance

**Affiliations:** School of Biological Sciences, Georgia Institute of Technology, Atlanta, Georgia, USA

**Keywords:** evolutionary genetics, health inequities, human genomics, natural selection, neutral evolution, polygenic diseases, ascertainment bias

## Abstract

**Background:** Although the prevalence of many complex diseases varies across human populations, the extent to which differences in the risks of common polygenic diseases are due to local adaptation is largely unknown. Focusing on the ten hereditary diseases with the largest global disease burden in terms of mortality rates, we leveraged GWAS findings from multiple ascertainment schemes to quantify collective signatures of selection acting on sets of disease-associated variants.

**Results:** First, we used PolyGraph to assess whether risk-associated loci have experienced directional shifts in allele frequencies. For each disease, these tests of polygenic adaptation revealed that most evolutionary branches did not exhibit any concerted change in predicted disease risks. Next, we developed a novel approach to quantify whether sets of disease-associated SNPs were enriched for outlier values from scans of selection compared to matched sets of control SNPs. Applying this approach to integrative haplotype scores, we did not observe strong signals of recent positive selection acting on common polygenic diseases. By contrast, application of our outlier approach to McVicker’s B statistics revealed that disease-associated SNPs are enriched for signatures of background selection. Furthermore, these tests of negative selection yielded consistent patterns regardless of whether disease-associated SNPs were ascertained in European, East Asian, or multi-ancestry cohorts.

**Conclusions:** While our results do not support a major role for recent positive selection or local adaptation in shaping population differences in polygenic disease risks, they do suggest that background selection continues to act on disease-associated loci across diverse human populations.

## Introduction

Disease risks have evolved substantially over recent human history [1, 2]. Increases in population size and changes in eating habits following the agricultural revolution led to an increase in nutritional and infectious diseases and a decline in the overall health of many ancient populations [3]. More recently the transition to modernity following the Industrial Revolution has resulted in increased rates of chronic non-communicable diseases [4, 5]. Indeed, the leading causes of death in sub-Saharan Africa have shifted from communicable diseases in children to non-communicable diseases in adults over the past three decades, with stroke, depression, diabetes, and ischemic heart disease dominating among middle-income countries [6]. The unique evolutionary histories experienced by different human populations can also cause disease risks to diverge.

Substantial heterogeneity in the mortality rates of non-communicable diseases exists across the globe, and these health inequities arise from a complex combination of socioeconomic, demographic, environmental, and genetic causes [7, 8] (Fig. 1). The narrow sense heritabilities of many complex diseases exceed 30% [9], and allele frequencies at disease-associated loci contribute to differences in hereditary disease risks. These population-level differences can be due to natural selection [10] or neutral processes like genetic drift and population bottlenecks [11, 12]. For example, individuals of African descent have higher rates of sickle cell disease due to evolutionary tradeoffs and selection against malaria [13]. By contrast, higher risks of cystic fibrosis in the Saguenay-Lac St. Jean region of Québec are due to founder effects [14]. However, unlike these two examples, many common diseases have complex non-Mendelian genetic architectures.

**Fig. 1.**
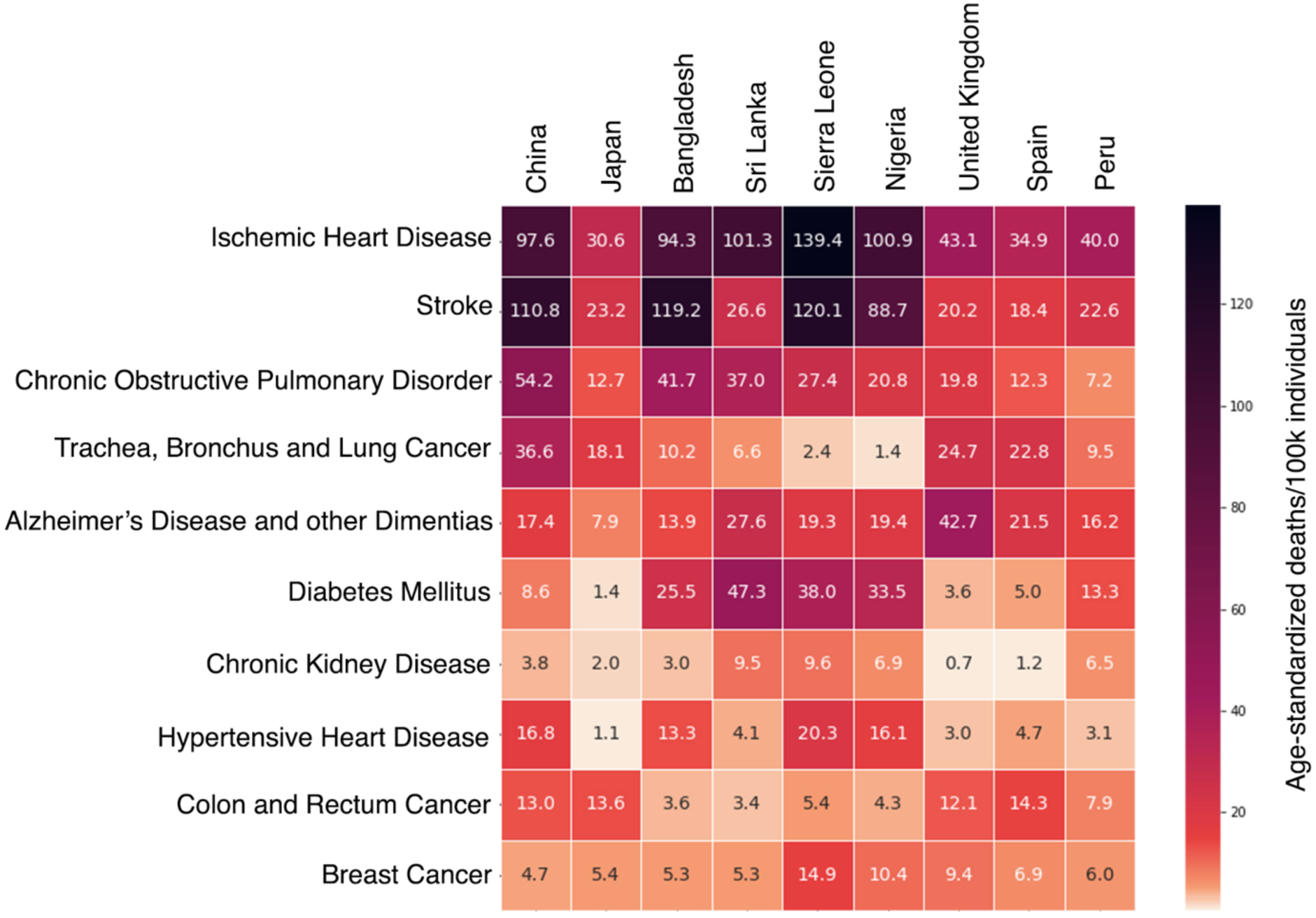
Heatmap of age-standardized mortality rates per 100,000 individuals for the 10 hereditary diseases with the largest global burden. Source of age-adjusted mortality rates: 2020 World Health Organization (WHO) Report [29]. Countries shown here have matching populations in the 1000 Genomes Project (1KGP).

One key insight gleaned from two decades of genome-wide association studies (GWAS) is that most non-communicable diseases are polygenic, i.e., hereditary disease risks are due to the cumulative effects of multiple SNPs [15–17]. These genetic architectures require polygenic tests of natural selection that focus on sets of trait-associated SNPs [18–20]. Analyses of ancient DNA have painted a mixed picture of human evolution during the Holocene (due in part to data quality issues and the need to correct for population continuity): some complex traits have appear to be evolving neutrally while others show evidence of polygenic selection [21–23]. Bayesian approaches examining the relationship between minor allele frequencies and the effect sizes of disease-associated SNPs have shown that negative and/or stabilizing selection widely impacts genetic variants that are associated with complex polygenic traits [24–26]. Similarly, recent analyses of conditional frequency spectra reveal that complex traits have experienced purifying and/or stabilizing selection [27]. Disease-associated SNP sets also exhibit signatures of evolutionary conservation over longer phylogenetic timescales [28]. One major limitation of most published studies of polygenic selection is that they focus on a single SNP set for each disease (most often from a European ancestry GWAS), and as such they do not control for ascertainment bias. Nor have published studies of polygenic selection focused on diseases with the largest global health burden. An open question is whether local adaptation and recent selection can explain why the mortality rates of these diseases vary substantially across populations or whether these differences are primarily due to socioeconomic factors (Fig. 1).

Here, we leveraged GWAS findings from European, East Asian and multi-ancestry cohorts to infer whether sets of disease-associated SNPs are enriched for signatures of natural selection (Table 1). We addressed three questions: 1) To what extent are population-level differences in hereditary disease burdens due to recent positive selection and polygenic adaptation? 2) Do complex diseases exhibit signatures of background selection? 3) Are our findings consistent across different GWAS ascertainment schemes and estimates of SNP effect sizes?

**Table 1.** Overview of the diseases, SNP sets, and polygenic tests of selection in this paper. Genetic ancestries and citations for each disease-associated SNP set are listed here, as are the number independent (r^2^ < 0.2) disease-associated SNPs. FDR-adjusted q-values from PolyGraph analyses are list here (as per Fig. 2). Percentile ranks refer to outlier enrichment comparisons between trait-associated SNP sets and 1000 matched control SNP sets. For each combination of disease and GWAS ascertainment scheme, the maximum integrated haplotype score (iHS) percentile rank is listed (as per Fig. 3), as is the relevant 1000 Genomes Project population code. Percentile ranks of McVicker’s B statistics quantify enrichment of signatures of background selection (as per Fig. 4). Alzheimer’s data were not available for East Asian and multi-ancestry GWAS. Additional details about each GWAS SNP set are described in Table S1.

| Disease | Ancestry of GWAS<br>SNP set [citation] | No. of SNPs<br>analyzed | PolyGraph<br>q-value | Max iHS %ile<br>(population) | McVicker's<br>B statistic %ile |
| --- | --- | --- | --- | --- | --- |
| Ischemic Heart<br>Disease | European [62] | 200 | 0.1689 | 98 (ITU) | 98.8 |
|  | East Asian [63] | 829 | 5.21x10<sup>-11</sup> | >99.9 (TSI) | >99.9 |
|  | Multi-ancestry [64] | 50 | 0.6884 | 86 (PEL) | 90.3 |
| Stroke | European [65] | 263 | 0.2951 | 88 (YRI) | 98.8 |
|  | East Asian [63] | 122 | 0.0941 | 84 (GWD) | 95.6 |
|  | Multi-ancestry [66] | 123 | 0.6884 | 74 (IBS) | 97.1 |
| COPD | European [67] | 79 | 0.9245 | 93 (PUR) | 98.8 |
|  | East Asian [63] | 92 | 0.7000 | 95 (JPT) | >99.9 |
|  | Multi-ancestry [68] | 100 | 0.6884 | 89 (GIH, PUR) | 99.8 |
| Lung Cancer | European [69] | 258 | 0.9245 | 92 (GBR) | 91.8 |
|  | East Asian [63] | 101 | 0.1144 | 92 (GBR) | 74.9 |
|  | Multi-ancestry [70] | 124 | 4.37x10<sup>-9</sup> | >99.99 (PEL) | 39.8 |
| Alzheimer's Disease | European [71] | 75 | 0.8876 | 94.5 (TSI) | 96.1 |
| Type 2 Diabetes | European [72] | 183 | 0.4697 | 85 (JPT) | 97.7 |
|  | East Asian [73] | 158 | 0.1777 | 98 (CLM) | 94.1 |
|  | Multi-ancestry [74] | 1661 | 0.6884 | 99 (PEL) | 99.9 |
| Chronic Kidney<br>Disease | European [75] | 2283 | 0.0113 | 98 (CDX, PEL) | >99.9 |
|  | East Asian [76] | 583 | 0.0010 | 57 (CHB) | >99.9 |
|  | Multi-ancestry [75] | 521 | 0.3186 | 70 (FIN, GWD) | >99.9 |
| Hypertensive Heart<br>Disorder | European [77] | 1124 | 0.0160 | 99 (PEL) | >99.9 |
|  | East Asian [76] | 315 | 0.0100 | 96 (ESN) | >99.9 |
|  | Multi-ancestry [78] | 226 | 0.6884 | 90 (CHS) | 99.9 |
| Colon Cancer | European [79] | 83 | 0.9245 | 78 (GIH) | 88.0 |
|  | East Asian [80] | 60 | 0.0061 | 72 (MSL) | 99.9 |
|  | Multi-ancestry [81] | 188 | 0.7225 | 62 (KHV) | 99.7 |
| Breast Cancer | European [82] | 242 | 0.9245 | 89 (CHS) | 99.2 |
|  | East Asian [63] | 118 | 0.0265 | 81 (CDX) | 78.6 |
|  | Multi-ancestry [83] | 100 | 0.9716 | 94 (CHS) | 98.9 |

## Results

Our tests of polygenic selection focused on the top 10 hereditary diseases with the largest public health burden as reported by the World Health Organization (WHO) [29]. These complex polygenic diseases included cardiometabolic diseases, (ischemic heart disease, hypertensive heart disease, stroke, type 2 diabetes), two chronic conditions (chronic obstructive pulmonary disease (COPD) and chronic kidney disease), three types of cancer (lung cancer, colon cancer, breast cancer), and Alzheimer’s disease (Fig. 1). For each disease we leveraged SNP sets from well-powered European, East Asian, and multi-ancestry GWAS cohorts (Table 1 and Table S1).

### Limited signatures of polygenic adaptation

First, we used PolyGraph to quantify signatures of polygenic adaption acting on sets of disease-associated SNPs. This approach leverages allele frequencies to identify directional shifts in predicted trait values along different evolutionary branches in an admixture graph [19]. FDR-adjusted q-values in Table 1 quantify whether a null hypothesis of neutral evolution can be rejected across an entire admixture graph. 20 of the 28 disease-GWAS ancestry combinations we analyzed did not show any statistically significant signature of polygenic adaptation. Focusing on SNP sets that were ascertained in European ancestry cohorts, we found that only two out of ten diseases listed in Table 1 have FDR-adjusted q-values of less than 0.05. Broadly similar patterns were observed for PolyGraph analyses that utilized SNP sets that were ascertained using East Asian or multi-ancestry GWAS cohorts (Table1). Five out of nine disease-associated SNP sets that were ascertained using East Asian GWAS had FDR-adjusted q-values less than 0.05 and only one out of nine disease-associated SNP sets that were ascertained using multi-ancestry GWAS cohort, had FDR-adjusted q-values that were less than 0.05. Moreover, most of the individual branches in our PolyGraph analyses do not exhibit statistically significant departures from neutrality (426 out of the 504 branch-specific *Q_B_* statistics listed in Table S2 have p-values > 0.05). Overall, the disease-associated SNP sets analyzed here appear to have experienced limited amounts of polygenic adaptation during recent human history.

Despite the general patterns described in the previous paragraph, some exceptions exist. PolyGraph results for disease-GWAS ancestry combinations that have FDR-adjusted q-values < 0.05 are shown in Fig. 2. Here, red shading indicates branches where risk-increasing alleles have increased in frequency, blue shading indicates branches were risk-increasing alleles have decreased in frequency, and gray shading indicates branches where there has been minimal directional change in the frequences of risk-increasing alleles. Despite nominal signatures of polygenic adaptation, the admixture graphs shown in Fig. 2 have a preponderance of gray branches. Furthermore, branch-specific signatures of polygenic adaptation for each disease appear to be sensitive to ascertainment bias, i.e. it matters whether disease-associated SNP sets come from European, East Asian, or multi-ancestry cohorts. For example, the branches driving the low FDR-adjusted q-values for chronic kidney disease and hypertensive heart disease vary for SNP sets that were ascertained using European or East Asian cohorts (compare panels A vs. D and panels B vs. E in Fig. 2). Similarly, only one lung cancer SNP set yields evidence of polygenic adaptation on the IBS_u branch (Table S2, p-values = 0.602, 0.562 and 0.00462 for European, East Asian, and multi-ancestry SNP sets, respective). Collectively, our PolyGraph analyses did not find a strong, replicable signature of polygenic adaptation on the diseases of interest.

**Fig. 2.**
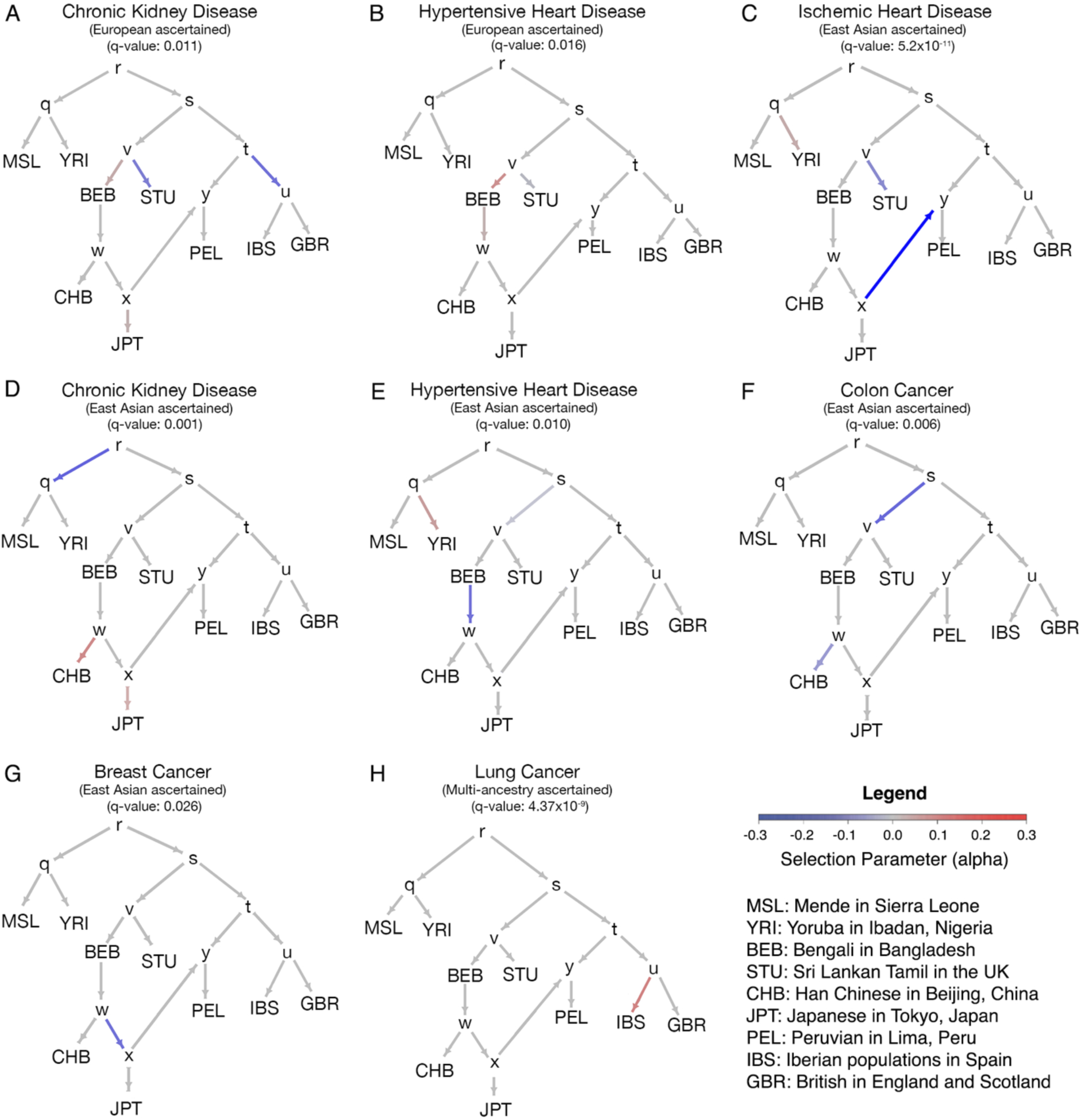
PolyGraph results do not show consistent evidence of polygenic adaptation across multiple GWAS ascertainment schemes. Note that 20 out of the 28 disease-GWAS ancestry combinations described in Table 1 did not show any evidence of polygenic selection – only the exceptions are plotted here. Allele frequencies from the 1KGP were used here, and population codes are indicated in the legend. q-values are FDR-adjusted. The selection parameter alpha reports the product of the selection coefficient for advantageous alleles and the duration of the selective process for each branch.

### Polygenic tests of outlier enrichment

Unlike PolyGraph, most other tests of selection focus on individual SNPs or genomic regions. These tests include integrative haplotype scores (iHS), which identify loci that have experienced recent positive selection [30], and McVicker’s B statistics, which identify loci under background selection [31]. Inferring the collective signatures of natural selection acting on polygenic diseases required the development of an outlier enrichment approach. Our novel approach leveraged effect size estimates from GWAS with and a kernel density estimation (KDE) framework to infer whether sets of disease-associated SNPs were more likely to contain outlier values of selection statistics compared to matched sets of control SNPs. These outlier enrichment analyses resulted in percentile ranks for each combination of disease, ascertainment scheme, and selection statistic, with higher percentiles indicating an excess of outlier values compared to control SNP sets. Additional details of our approach can be found in the Methods section.

### Sparse signatures of recent positive selection

To identify diseases under recent positive selection, we employed our outlier enrichment approach to integrated Haplotype Score (iHS) statistics from the 1000 Genomes Project (1KGP) [32]. Note that this approach is complementary to PolyGraph, as iHS statistics quantify extended haplotype homozygosity within each population while PolyGraph relies on allele frequency differences between populations. In total, 26 global populations were analyzed for 10 diseases and three ascertainment schemes (apart from Alzheimer’s disease which lacked East Asian and multi-ancestry SNP sets).

Overall, we found that disease-associated SNP sets were not enriched for iHS outliers when compared to control SNP sets (Fig. 3 and Table S3). Focusing on European ascertained GWASs, most diseases show low percentile values in all 26 populations, i.e. they were not enriched for signatures of recent positive selection. Only six out of 260 tests had percentile ranks above 95 (p-value = 0.991, one-tailed binomial test with alpha = 0.05). Exceptions include ischemic heart disease in South Asian populations, namely Bengalis in Bangladesh (BEB), Indian Telugu in the UK (ITU), and Sri Lankan Tamil in the UK (STU). The admixed population from Lima, Peru (PEL) showed enrichment for high iHS statistics for chronic kidney disease and hypertensive heart disorder. However, when the same diseases were tested without considering effect sizes (i.e., all SNPs were given equal weight in the KDE), the iHS outlier enrichments disappeared (Table S4 and Fig. S1). This supports the notion that most disease-associated SNP sets are not under strong positive selection, and that potential exceptions may be due to a small number of lead SNPs with large effect sizes.

**Fig. 3.**
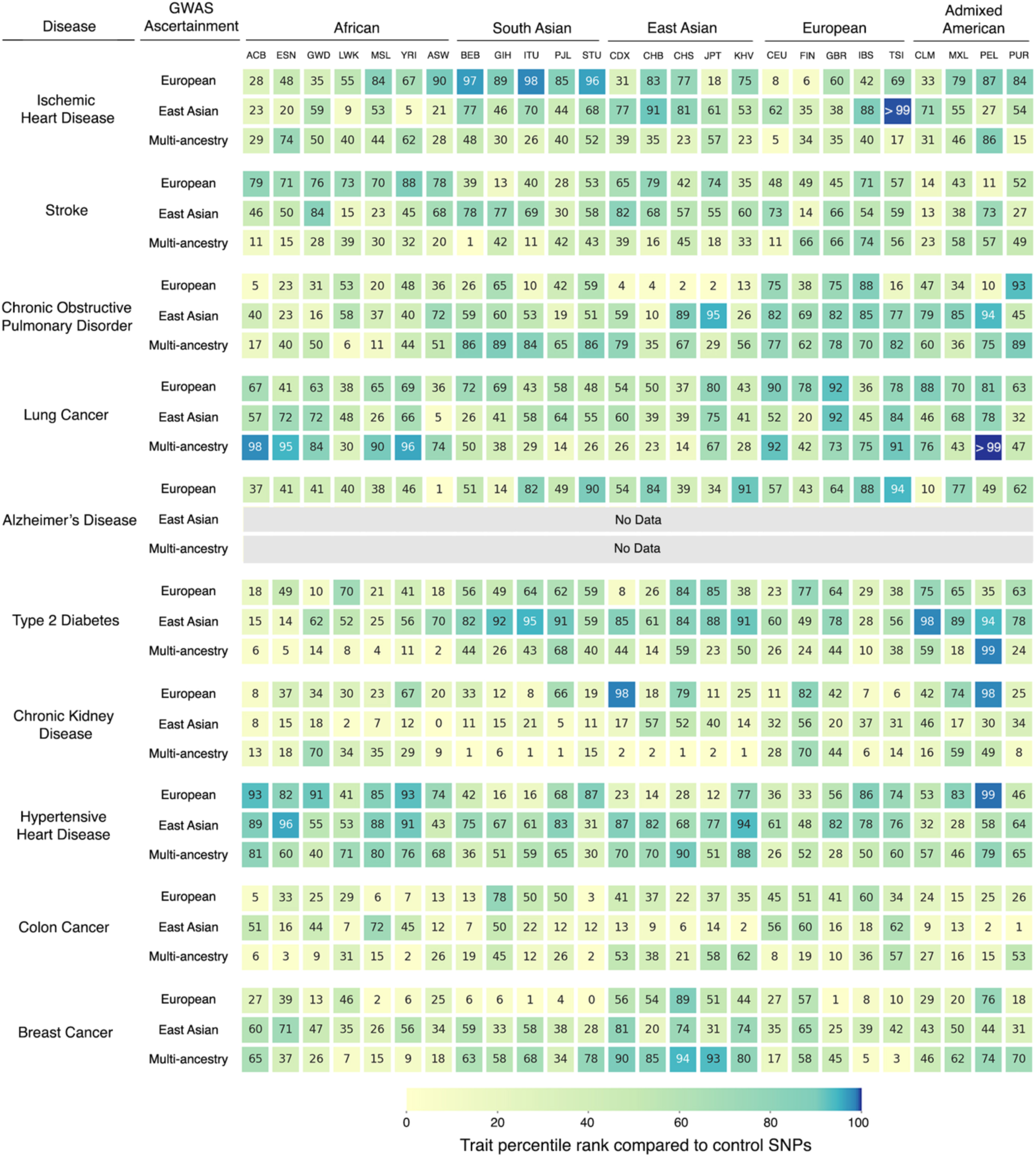
Sparse signals of recent positive selection acting on complex diseases. Plotted here are results from SNPs sets that were ascertained in European, East Asian, and multi-ancestry GWAS. Percentile ranks quantify how much disease-associated loci are enriched for outlier values of integrative haplotype scores (|iHS| >1.96) compared to 1000 sets of control SNPs, with darker shading indicating stronger polygenic signatures of recent positive selection. Population codes, i.e., column headings, are from the 1000 Genomes Project (1KGP).

We observed similar results for disease associations that were ascertained in East Asian and multi-ancestry cohorts. Only five out of 234 tests of East Asian-ascertained SNP sets showed strong enrichments for recent positive selection (p-value = 0.992, one-tailed binomial test with alpha = 0.05). Similarly, only five out of 234 tests of that were ascertained using multi-ancestry SNP sets also showed strong enrichments for recent positive selection (p-value = 0.992, one-tailed binomial test with alpha = 0.05). Importantly, none of the high iHS percentile exceptions replicate across all three SNP ascertainment schemes.

### Evidence of background selection

Although we found only minimal signatures of polygenic adaptation and recent positive selection, disease-associated loci may be subject to other types of selection, including background selection (the reduction of genetic diversity at a locus due to negative selection against linked deleterious alleles). With this in mind, we used McVicker’s B statistics [31] and our outlier enrichment approach to measure whether sets of disease-associated loci were enriched for signatures of background selection. Fig. 4 shows the percentile rank for each set of disease-associated SNPs compared to matched control sets for three different GWAS ascertainment schemes (see Table 1 for exact values). SNP sets that were ascertained in European ancestry populations yielded percentile ranks that ranged from 88.0 (colon cancer) to above 99.9 (chronic kidney disease and hypertensive heart disease), indicating that disease-associated SNPs are more likely to have outlier values of McVicker’s B statistics compared to matched controls. Eight out of ten diseases had percentile ranks above the 95^th^ percentile of matched control sets, a statistically significant fraction (p-value = 1.61 x 10^-9^, one-tailed binomial test with alpha = 0.05). We see similar patterns for disease-associated SNP sets that were ascertained in East Asian populations: six out of nine diseases have percentile ranks above 95 (p-value = 1.15 x 10^-6^, one-tailed binomial test with alpha = 0.05). Likewise, seven out of nine diseases have percentile ranks over 95 when SNP sets that were ascertained in multi-ancestry GWAS were analyzed (p-value = 2.57 x 10^-8^, one-tailed binomial test with alpha = 0.05). Overall, we found that disease-associated SNPs were strongly enriched for signatures of background selection regardless of whether the disease-associated loci were ascertained using a European, East Asian, or multi-ancestry cohort.

**Fig. 4.**
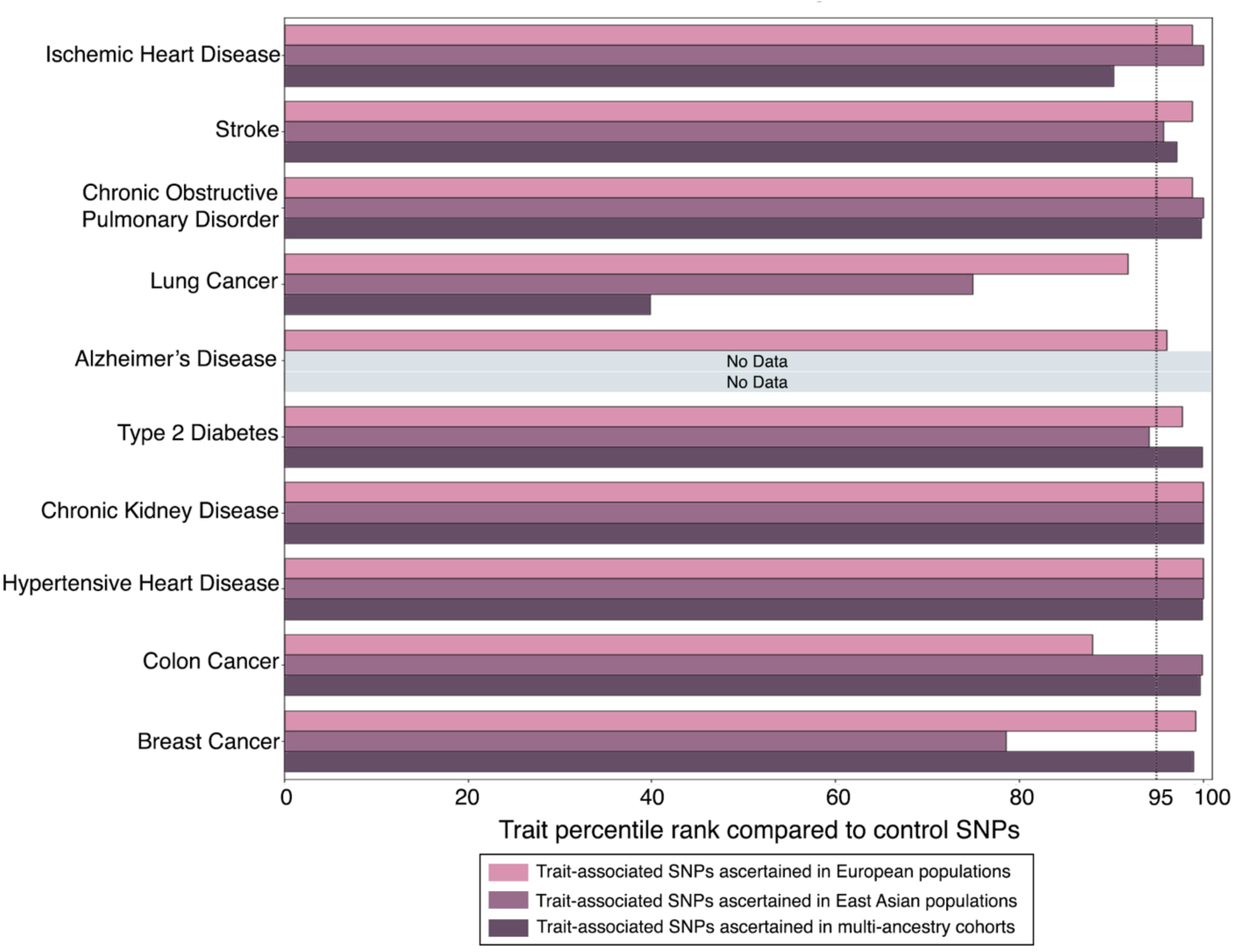
Disease-associated SNP sets are enriched for signatures background selection. Results from SNP sets that were ascertained in European, East Asian, and multi-ancestry GWAS are plotted here (exact percentile ranks can be found in Table 1). Percentile ranks for each disease involve comparison between sets of disease-associated to 1000 sets of control SNPs. The dotted line marks the 95^th^ percentile of control SNP sets. As per [60], a McVicker’s B statistic outlier threshold of 0.317 was used.

Stroke, COPD, chronic kidney disease, and hypertensive heart disease showed the strongest enrichment for background selection (percentile rank above 99.9 for all ascertainment schemes). By contrast, lung cancer SNP sets did not show any significant enrichment for signatures of background selection, perhaps due to a lack of historical selection pressures against smoking during most of human history [33]. To ensure the robustness of our findings, we tested whether our results were consistent across different outlier thresholds of McVicker’s B (Table S5 and Fig. S2). Once again, SNPs associated with diseases that have a large global health burden showed enrichment for signatures of background selection, though this trend was weaker for more stringent outlier thresholds. We further tested the consistency of our background selection results by not weighting the KDE by effect size (Table S6 and Fig. S3). Focusing on European-ascertained disease associations and giving all SNPs in each SNP set equal weight, we found that nine out of ten diseases were still enriched for signatures of background selection (p-value = 1.87 x 10^-11^, one-tailed binomial test with alpha = 0.05).

## Discussion

Focusing on the ten diseases with the largest global health burden, we tested whether sets of disease-associated SNPs are enriched for signatures of natural selection. PolyGraph analyses and outlier enrichment tests of iHS statistics revealed minimal evidence for polygenic adaptation and recent positive selection acting on the diseases studied here. By contrast, outlier enrichment tests of McVicker’s B statistics revealed that disease-associated SNPs tend to be found in genomic regions that have lower amounts of genetic diversity, even after controlling for allele frequency and distance to the nearest exon. While SNP sets that are associated with polygenic diseases show limited signatures of local adaptation, our results reinforce the role of background selection as a long-standing evolutionary influence on disease-associated loci across human lineages.

SNP ascertainment bias presents a major challenge when using GWAS data [34, 35], and the ability to detect genetic associations depends on both allele frequencies and effect sizes in the discovery population [36]. One byproduct of this is that sets of disease-associated SNPs can vary across studies, particularly when the genetic ancestries of study participants differ. When possible, we analyzed three different ascertainment schemes for each disease, i.e., SNP sets that were ascertained in European, East Asian, and multi-ancestry cohorts (Table 1). While some weak signals of polygenic adaptation were identified, these findings were not replicated across different ascertainment schemes.

We note that the diseases studied here are highly polygenic, and although they might not show strong selection signatures at the overall trait level, individual SNPs can still be under appreciable selection pressures [37, 38]. This point is magnified for species with small historic effective population sizes like our own, and for diseases with late ages of onset [39–41]. Disease-associated alleles can also affect multiple traits. Because of this, antagonistic pleiotropy could in principle dilute directional signals of selection [42]. Furthermore, our study focused on common variants associated with each disease, and future studies will benefit from examining the impact of rare variants on the population-specific genetic architectures of complex diseases [43, 44]. Additionally, future studies that combine genetic and environmental data will provide deeper insights into the risks of common complex diseases and why they differ across diverse human populations [45, 46].

### Conclusions

One implication of our study is that polygenic risk score (PRS) differences between populations should not be interpreted as prima facia evidence of local selection. Differences in (predicted) hereditary disease risks across populations may well be due to genetic drift [47, 48] and/or ascertainment bias [34]. Although our findings do not point to local adaptation as a primary driver of health inequities, they do suggest that background selection continues to act on disease-associated loci across diverse human populations.

## Methods

### Processing of GWAS results

To investigate the evolutionary genetics of these complex polygenic diseases, we extracted GWAS summary statistics from ten diseases across three distinct types of study cohorts: European, East Asian, and multi-ancestry (Table 1). Due to an insufficient number of significant associations identified for Alzheimer’s Disease in East Asian and multi-ancestry ascertained GWAS, this trait was excluded from our ascertainment bias testing. GWAS sample sizes exceeded 72,000 individuals for each combination and disease and study cohort (Table S1). Marginally significant autosomal SNPs with a p-value < 5×10^-5^ were extracted from each GWAS. Subsequently, LD clumping was performed to isolate independent associations with an r^2^ < 0.2 within the respective ascertained population, utilizing Plink 1.9 [49] and 1KGP phase 3 data [32] as a reference. In the case of multi-ancestry GWASs, clumping was performed separately in European, East Asian, and African reference populations from the 1KGP, and an intersection of independent associations across populations were used for further analysis (Details of SNPs pre- and post-clumping can be found in Table S1. To ensure uniformity, the LiftOver tool [50] was employed to convert coordinates of all GWAS SNPs to the GRCh37 assembly (hg19 build) of the human reference genome. When these analyses were conducted well-powered GWAS results for Alzheimer’s disease were not available for East Asian or multi-ancestry cohorts.

### Generating control sets of SNPs

In all our analyses, autosomal control SNPs were obtained using SNPSnap [51]. Each trait associated SNP had approximately 5000 matched independent (r^2^ < 0.2 in respective population) SNPs. Matching criteria included allele frequency (+/- 5%), number of SNPs in LD (r^2^ < 0.5) with the target SNPs (+/- 10%), gene density (+/- 10%) in the ascertained population, and distance to the nearest exon (+/- 5%). This latter filter is particularly important, as simply selecting control SNPs based on distance to nearest gene would treat intronic SNPs the same as exonic SNPs. SNPSnap automatically removes any SNPs within the HLA region when matching for control SNPs. For European and East Asian ascertained GWAS, controls SNPs were matched within their respective populations from the 1KGP. In the case of multi-ancestry studies, control SNPs were matched across pooled data from European, East Asian, and African populations. Lists of GWAS and matched sets of control SNPs are available on GitHub (see Data Availability). We note that the top GWAS hits for a trait can often capture a sufficient fraction of the total SNP heritability of polygenic traits, and there are diminishing returns for polygenic scores that incorporate more than the top few hundred independent hits [52–54].

### PolyGraph analyses (polygenic adaptation)

To investigate signals of polygenic adaptation, we used PolyGraph [19], a Markov Chain Monte Carlo (MCMC) algorithm that leverages admixture graph information to detect traces of polygenic adaptation in populations. PolyGraph detects adaptation of polygenic traits due to allele frequency shifts at multiple loci using an admixture graph framework that considers the historical divergence of populations. It uses the ancestral and derived allele frequencies for each disease-associated SNP for every population along with effect sizes and compares them to control distributions. In contrast to iHS statistics, which are ideal for detecting hard sweeps, PolyGraph analyses are able to detect soft sweeps from standing genetic variation.

We used control SNPs from the enrichment analyses to build an admixture graph with *MixMapper* v2.0 [55]. In the original PolyGraph paper, the authors caution against using programs like *TreeMix* [56], which scale drift values by the heterozygosity of ancestral nodes, as this could alter the interpretation of results from PolyGraph [19]. Although *MixMapper* and *TreeMix* give similar topologies, we used *MixMapper* to build the admixture graph to ensure accurate results. We made scaffold trees with eight continental populations and added the population from Peru (PEL) as an admixed population. We ran PolyGraph using 1,000,000 MCMC steps. PolyGraph reported a selection parameter alpha for each disease, a product of the selection coefficient for the advantageous allele and the duration of the selective process, and a p-value for selection on the entire admixture graph. To correct for multiple testing, we calculated FDR-adjusted q-values from the overall p-values of selection from PolyGraph.

### Trait-level distributions of selection statistics

To integrate SNP-level information from test statistics into a comprehensive trait-level distribution, we employed kernel density estimation (KDE). This method allowed us to derive a probability distribution of the test statistic for each trait. Unlike traditional estimation techniques, KDE is a nonparametric approach that does not assume that the data follows a known distribution. In our implementation, we opted for a Gaussian kernel and conducted a five-fold cross-validation using GridSearchCV [57] to determine the optimal kernel bandwidth for the KDE. Since each associated SNP also has an effect size or beta, we also weighted the SNPs according to their absolute effect sizes. Thus, outliers count more for disease-associated SNPs that have larger effect sizes. Finally, we obtained a probability density function of the test statistic for each set of disease-associated SNPs from the KDE.

For each set of disease-associated SNPs, we obtained 1000 sets of matched control SNPs matched for allele frequency, linkage disequilibrium (LD) patterns in the ascertained populations, gene density, and distance to the nearest exon. For each SNP set, we identified the proportion of SNPs, weighted by effect size, that exceeded standard outlier thresholds (see below). Enrichment tests involved comparing outlier proportions of disease-associated SNP sets to control sets to generate a percentile rank, with higher percentiles indicating greater trait-level signatures of selection (Fig. S4). Our approach differed from that of other research teams [28] in that we looked for outlier enrichment, as opposed to trait averages, plus we weighted each SNP by effect size.

### Outlier enrichment of iHS (recent positive selection)

We used iHS to measure recent positive selection. This statistic compares the haplotype homozygosity surrounding an ancestral allele to that of the derived allele at the same position. iHS values are normalized, with iHS > 1.96 indicating that haplotypes with the ancestral allele are longer and positively selected than those with the derived alleles, while iHS < 1.96 indicates that the derived allele has longer haplotypes. Therefore, both extreme positive and negative iHS values (|iHS| > 1.96) were considered indicative of selection [58]. Our analyses used pre-computed values of iHS statistics in 26 global populations from the 1KGP [30].

To detect trait-level signatures of positive selection, we used the probability density function for iHD to calculate the weighted proportion of trait-associated variants with |iHS| > 1.96. Here, we used area under the curve (AUC) statistics to quantify the proportion of a SNP set with outlier values of iHS values. Our test of enrichment involved generating a percentile rank for the trait by comparing the iHS outlier AUC for the trait-associated SNP set to 1000 AUC values for matched control sets of SNPs. Our test of enrichment involved generating a percentile rank for the trait by comparing outlier AUC statistics for the trait-associated SNP set to 1000 matched outlier AUC statistics for matched control sets of SNPs (Fig. S4). High percentile ranks signify that trait-associated SNPs are enriched for extreme iHS values. We also tested whether our recent positive selection results were robust to ignoring effective size differences – focusing on European ancestry GWAS hits (Table S4 and Fig. S1). This involved modifying the iHS KDE by using the same effect size for every SNP, i.e. by weighting each SNP equally.

### Outlier enrichment of McVicker’s B (background selection)

We used McVicker’s B statistic as a measure of background selection [31]. McVicker’s B values that are close to 0 indicate nearly complete removal of diversity due to background selection and values close to 1 indicate minimal impact. Using BEDTools [59], we extracted McVicker’s B values for GWAS SNPs and their matched controls. Previous research suggests that McVicker’s B ≤ 0.317 is a threshold for the lowest 5% of McVicker’s B values across the human genome [60]. Using the probability density function for McVicker’s B, we calculated the weighted proportion of trait-associated variants with McVicker’s B ≤ 0.317. Here, we used AUC statistics to quantify the proportion of a SNP set with outlier values of McVicker’s B. Our test of enrichment involved generating a percentile rank for the trait by comparing the McVicker’s B outlier AUC for the trait-associated SNP set to 1000 AUC values for matched control sets of SNPs (Fig. S4). High percentile ranks signify that trait-associated SNPs are enriched for extreme McVicker’s B statistics. Previous research has demonstrated that McVicker’s B statistic reliably preserves the correct rank order of SNPs [60, 61]. Nevertheless, to ensure the robustness of our findings, we verified our results are consistent across more stringent thresholds (McVicker’s B ≤ 0.2 and 0.1), focusing European ancestry GWAS hits (Table S5 and Fig. S2). We also tested whether our background selection results were robust to ignoring effective size differences, once again focusing on European ancestry GWAS hits (Table S6 and Fig. S3). This involved modifying the McVicker’s B KDE by using the same effect size for every SNP, i.e. by weighting each SNP equally.

## Data Availability

All data produced in the present work are contained in the manuscript.

## Declarations

### Ethics approval

All the GWAS summary statistic data analyzed in this paper was publicly available.

### Consent for publication

Not applicable

### Availability of data and materials

The GWAS summary statistics used in this paper are publicly available. Details about specific studies and SNPs used for analyses can be found in S1 Table and they are available via GitHub at https://github.com/LachanceLab/polygenic_selection. Custom code that was used to quantify polygenic signatures of natural selection is also available on GitHub at https://github.com/LachanceLab/polygenic_selection.

### Author contributions

UH and JL conceived this study and developed the study methodology. UH curated GWAS datasets, conducted polygenic tests of selection, and performed data visualization. JL supervised this research and provided funding. UH and JL co-wrote and edited this manuscript.

## Acknowledgements

We thank Rohini Janivara, Aaron Pfennig, Mimi Holness, Ianne Lauder, King Jordan, Greg Gibson and members of the Center for Integrative Genomics at Georgia Institute of Technology for their insight and helpful comments.

## Additional files

### Additional file 1

Tables S1-S6 are embedded in a tabbed .xslx file. **Table S1**: Additional details about GWAS SNP sets. Details include citations, the genetic ancestry of the each GWAS study cohort, sample sizes, number of SNPs before and after LD clumping, and links to SNP lists (including iHS statistics for each SNP). **Table S2**: Branch statistics for PolyGraph analyses. Summary statistics are shown for ten complex diseases and three different GWAS ascertainment schemes: European, East Asian, and multi-ancestry cohorts. Branch-specific and admixture graph-wide statistics are included. **Table S3**: Exact iHS outlier enrichment percentile rank statistics for multiple ascertainment schemes and 26 global populations from the 1KGP. Higher percentile ranks indicate enrichment for signatures of recent positive selection. **Table S4**: Effects of weighting the KDE by effect size for iHS statistics. Outlier enrichment percentile ranks are for European ascertained SNP sets.Higher percentile ranks indicate enrichment for signatures of recent positive selection. **Table S5**: Comparisons of different outlier thresholds for McVicker’s B statistics. Outlier enrichment percentile ranks are for European ascertained SNP sets. Higher percentile ranks indicate enrichment for signatures of background selection. **Table S6**: Effects of weighting the KDE by effect size for McVicker’s B statistics. Outlier enrichment percentile ranks are for European ascertained SNP sets. Higher percentile ranks indicate enrichment for signatures of background selection.

### Additional file 2

Figures S1-S4 are combined in a singled merged file.

