## Supplementary material for "The evolutionary genetics of polygenic diseases with the largest global burden in mortality rates": Figures S1-S4

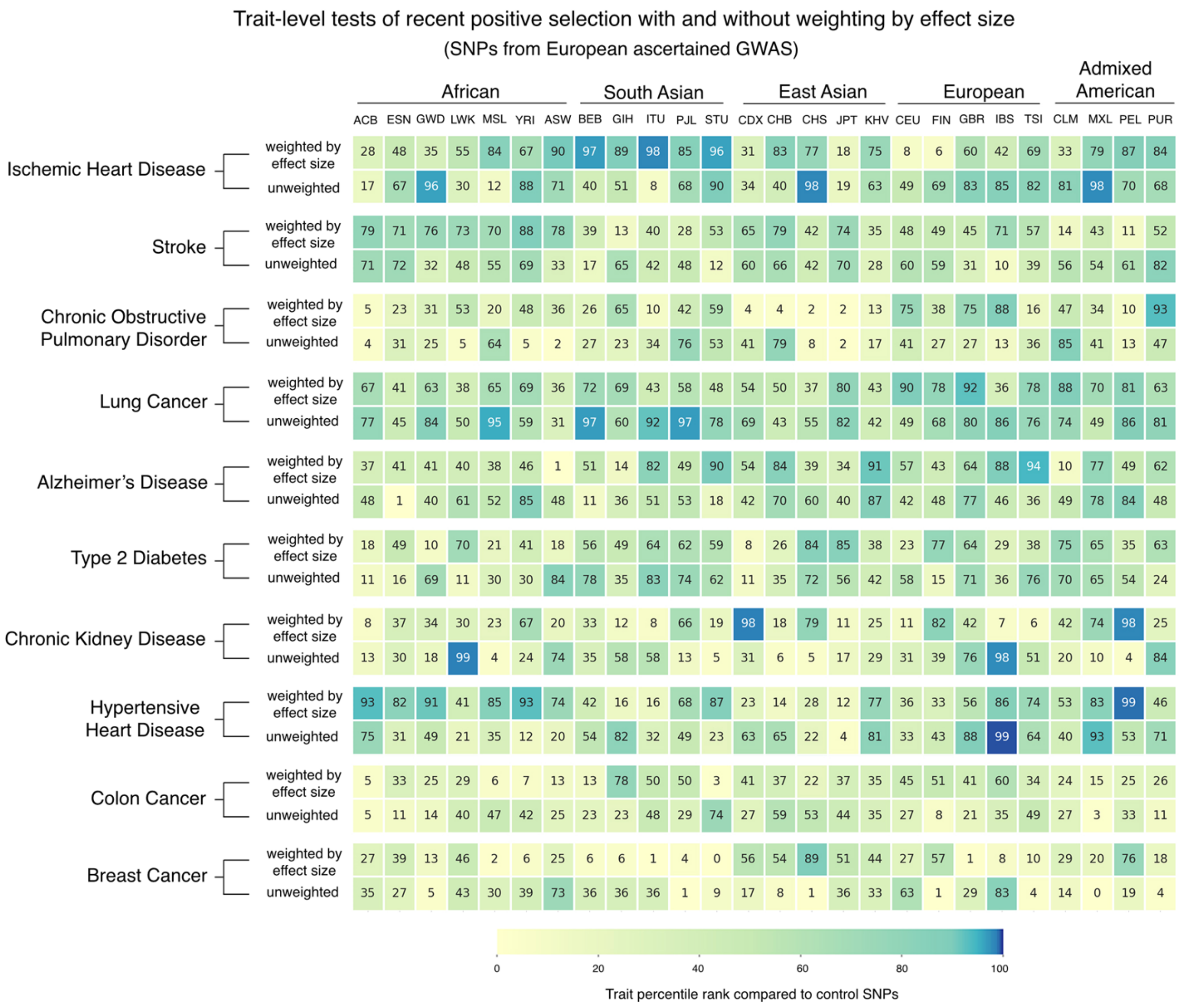


**Fig. S1.** Outlier enrichment tests of iHS with and without considering GWAS effect sizes. Plotted here are results from SNPs sets that were ascertained using European ancestry GWAS. Percentile ranks quantify how much disease-associated loci are enriched for outlier values of integrative haplotype scores (|iHS| >1.96) compared to 1000 sets of control SNPs, with darker shading indicating stronger polygenic signatures of recent positive selection. Exact percentile ranks can be found in Table S4. Population codes, i.e., column headings, are from the 1000 Genomes Project (1KGP). Weighted KDE percentiles use absolute values of effect sizes from GWAS, while unweighted KDE percentiles assume that every disease-associated SNP in a SNP set has the same effect size.

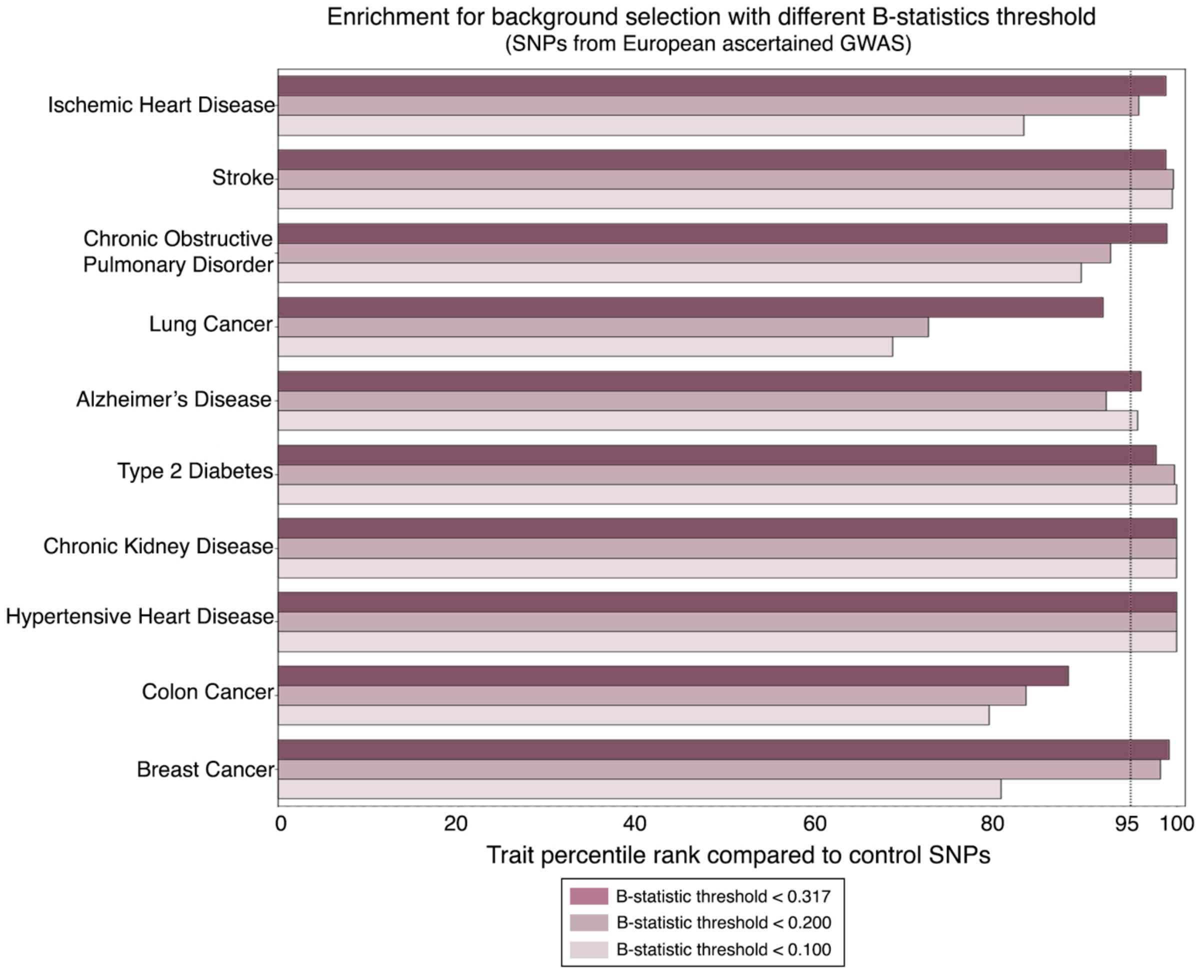


**Fig. S2.** Outlier enrichment tests of McVicker's B statistics with different thresholds. Results from SNP sets that were ascertained in European ancestry GWAS are plotted here (exact percentile ranks can be found in Table 1). Percentile ranks for each disease involve comparison between sets of disease-associated to 1000 sets of control SNPs. The dotted line marks the 95$^{th}$ percentile of control SNP sets. Here we explored three different outlier thresholds for McVicker's B: 0.317, 0.200, and 0.100.

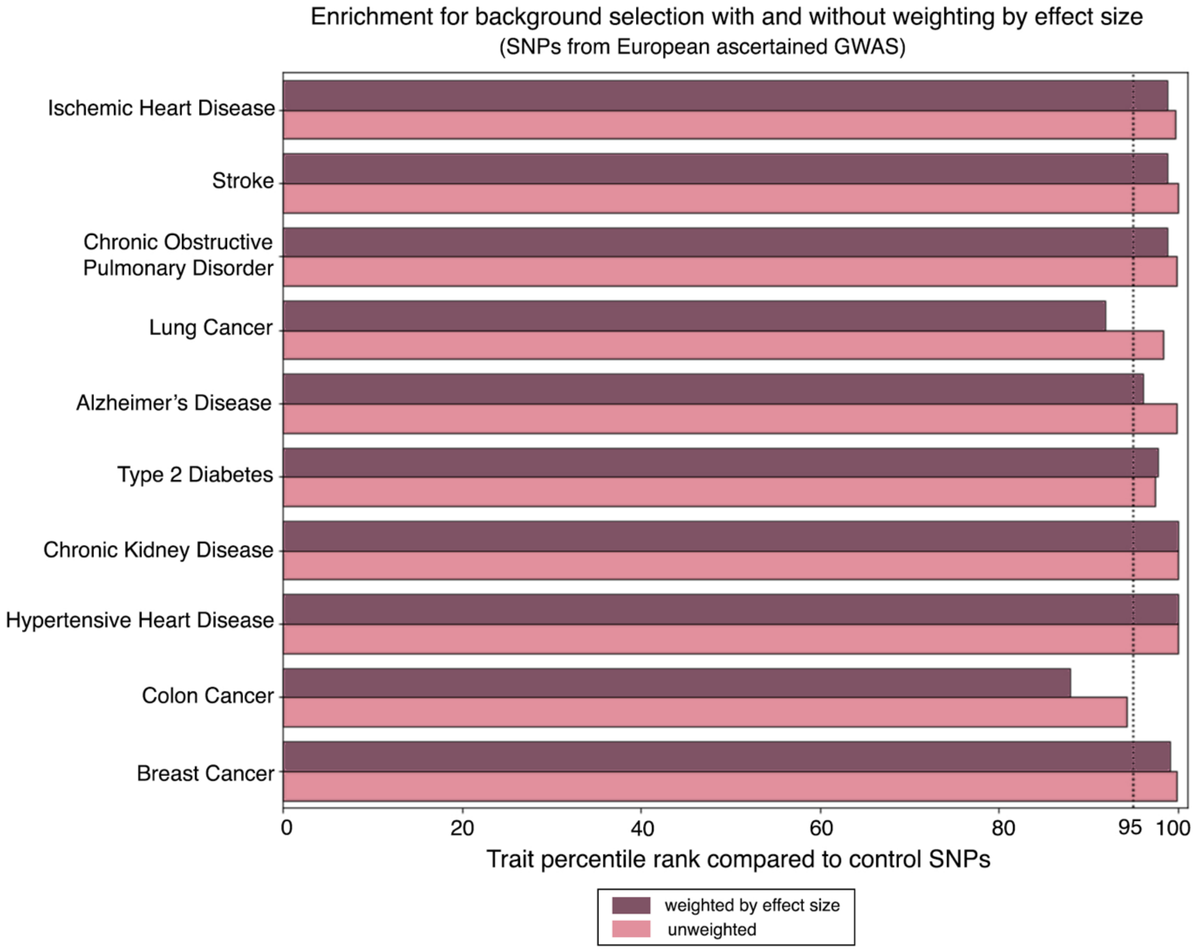


**Fig. S3.** Outlier enrichment tests McVicker's B statistics with and without considering GWAS effect size. Results from SNP sets that were ascertained in European ancestry GWAS are plotted here (exact percentile ranks can be found in Table S6). Percentile ranks for each disease involve comparison between sets of disease-associated to 1000 sets of control SNPs. The dotted line marks the 95$^{th}$ percentile of control SNP sets. As per [60], a McVicker's B statistic outlier threshold of 0.317 was used. Weighted KDE percentiles use absolute values of effect sizes from GWAS, while unweighted KDE percentiles assume that every disease-associated SNP in a SNP set has the same effect size.

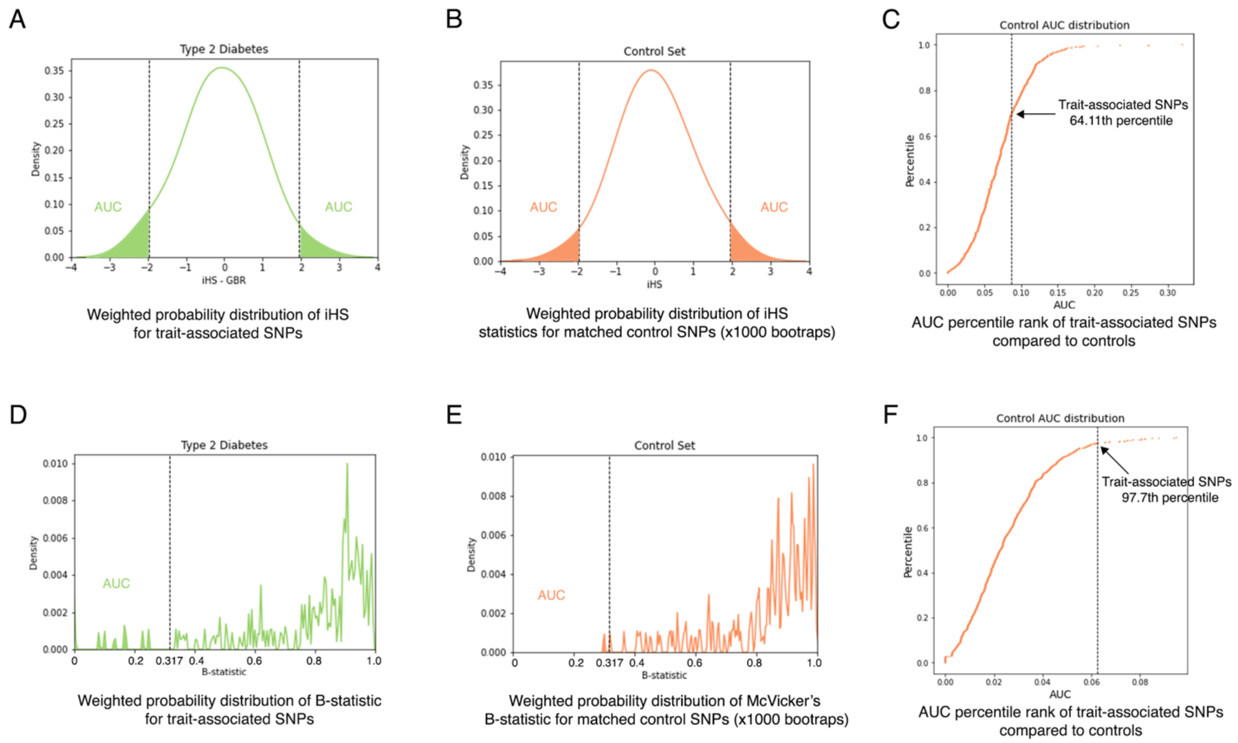


**Fig. S4.** Schematic illustrating our outlier enrichment approach. Panel A: KDE of iHS statistics for a set of disease-associated SNPs. Outlier threshold: iHS > |1.96|. Panel B: KDE of iHS statistics for a represented set of matched control SNPs. Panel C: iHS percentile rank inferred from comparisons of the the outlier area under the curve (AUC) for a set of disease-associated SNPs to AUC from 1000 control sets of SNPs. Panel D: KDE of McVicker's B statistics for a set of disease-associated SNPs. Outlier threshold: McVicker's B < 0.317. Panel E: KDE of McVicker's B statistics for a represented set of matched control SNPs. Panel F: McVicker's B statistic percentile rank inferred from comparisons of AUC for a set of disease-associated SNPs to AUC from 1000 control sets of SNPs. All panels use Type 2 Diabetes SNPs and GBR data from the 1KGP.
